# Evaluating the trade-off between transmissibility and virulence of SARS-CoV-2 by mathematical modeling

**DOI:** 10.1101/2021.02.27.21252592

**Authors:** Hyun Mo Yang, Luis Pedro Lombardi Junior, Ariana Campos Yang

## Abstract

**Background:** At the beginning of 2020, SARS-CoV-2 spread to all continents, and since then, mutations have appeared in different regions of the world. The appearance of more virulent mutations leads to asseverate that they are also more transmissible. We analyzed the lower and higher virulent SARS-CoV-2 epidemics to establish a relationship between transmissibility and virulence based on a mathematical model.

**Methods:** A compartmental mathematical model based on the CoViD-19 natural history encompassing the age-dependent fatality was applied to evaluate the SARS-CoV-2 transmissibility and virulence. The transmissibility was measured by the basic reproduction number *R*_0_ and the virulence by the proportion of asymptomatic individuals. The model parameters were fitted considering the observed data from São Paulo State.

**Results:** The numbers of severe CoViD-19 and deaths are three times higher, but *R*_0_ is 25% lower in more virulent SARS-CoV-2 transmission than in a less virulent one. However, the number of more virulent SARS-CoV-2 transmitting individuals is 25% lower, mainly due to symptomatic individuals’ isolation, explaining the increased transmission in lower virulence.

**Conclusions:** The quarantine study in São Paulo State showed that the more virulent SARS-CoV-2 resulted in a higher number of fatalities but less transmissible than the less virulent one. One possible explanation for the number of deaths surpassing that predicted by the low virulent SARS-CoV-2 infection could be the transmission of more virulent variant(s).

## 1 Introduction

The severe acute respiratory syndrome coronavirus 2 (SARS-CoV-2), an RNA virus, can be transmitted by droplets that escape lungs through coughing or sneezing and infects humans (direct transmission), or is deposited in surfaces and infects humans when in contact with this contaminated surface (indirect transmission) [1] [2]. Coronavirus disease 2019 (CoViD-19) caused by SARS-CoV-2 infection was declared a pandemic by the World Health Organization on March 11, 2020. In general, the fatality rate in elder patients (60 years or more) is much higher than those with 60 years or less [3].

Since the worldwide outbreak of CoViD-19, like all RNA-based viruses, SARS-CoV-2 mutates faster but tends to change more slowly than HIV or influenza viruses [4]. “A virus with one or several new mutations (small changes during the replication) is referred to as a variant of the original virus” [4]. Currently, three variants challenge the vaccination’s control efforts – B.1.1.7 (United Kingdom), B.1.351 (South Africa), and P.1 (Brazil) (see [5] and references therein). Many countries closed their frontiers, mostly travelers from United Kingdom, South Africa, and Brazil, to control these variants. It is currently accepted that the more virulent variants of SARS-CoV-2 are also more transmissible based on the increased deaths caused by these strains [6].

Based on a mathematical model, we evaluate the trade-off between transmissibility and virulence. We hypothesize that the transmissibility is measured by the transmission rate (ultimately, by the basic reproduction number *R*_0_), and the virulence is assessed by the ratio between asymptomatic and symptomatic individuals. Using the data collected from São Paulo State (Brazil), we estimate the basic reproduction number considering two virulence levels. We take as our framework the CoViD-19 epidemic in São Paulo State to evaluate the trade-off between transmissibility and virulence.

## 2 Material and methods

The SARS-CoV-2 transmission model is described by the system of ordinary differential equations presented in Appendix A, named the SQEAPMDR model. We drop out the pulses in equation (A.2) in Appendix A and transfer them to the boundary conditions. Hence, equations for susceptible individuals are

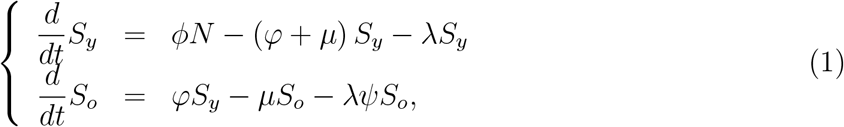

for isolated individuals *Q*_*j*_ and infected individuals, with *j* = *y, o*,

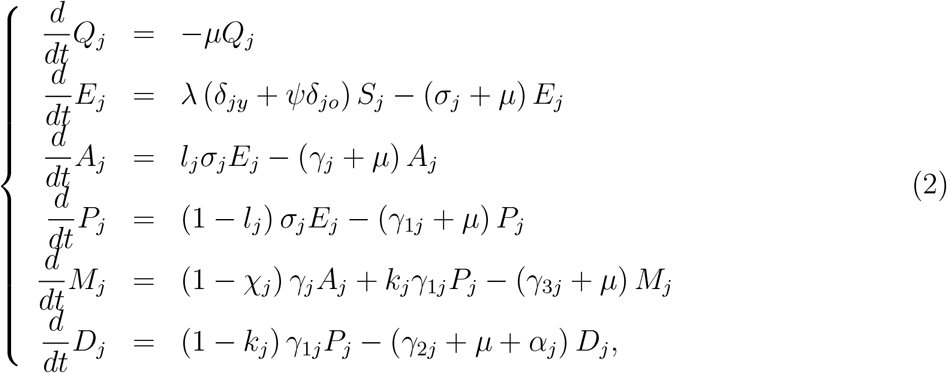

and for recovered individuals,

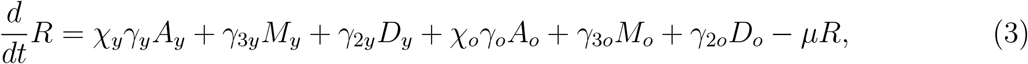

where *N*_*j*_ = *S*_*j*_ + *Q*_*j*_ + *E*_*j*_ + *A*_*j*_ + *P*_*j*_ + *M*_*j*_ + *D*_*j*_, and *N* = *N*_*y*_ + *N*_*o*_ + *I* obeys

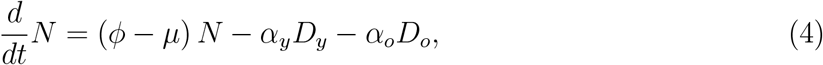

with the initial number of population being *N* (0) = *N*_0_ = *N*_0*y*_ + *N*_0*o*_, where *N*_0*y*_ and *N*_0*o*_ are the size of young and elder subpopulations at *t* = 0. Table 1 summarizes the model compartments (variables).

**Table 1:** Summary of the model variables, for *j* = *y* (young) and *j* = *o* (elder).

| Symbol | Meaning |
| --- | --- |
| $S_j$ | Susceptible individuals |
| $Q_j$ | Isolated individuals |
| $E_j$ | Exposed and incubating SARS-CoV-2 individuals |
| $A_j$ | Asymptomatic individuals |
| $P_j$ | Pre-diseased (pre-symptomatic) individuals |
| $M_j$ | Mild (non-hospitalized) CoViD-19 individuals |
| $D_j$ | Severe (hospitalized) CoViD-19 individuals |
| $R$ | Recovered (immune) individuals |

The initial and boundary conditions supplied to the system of equations (1), (2) and (3) are given by equations (A.3) and (A.5) in Appendix A.

Table 2 summarizes the model parameters. The description of the assigned values to the CoViD-19 natural history parameters can be found in [7]. The values of the parameters not shown in the table are estimated considering two SARS-CoV-2 virulence levels.

**Table 2:** Summary of the model parameters (*j* = *y, o*) and values (rates in *days*^−1^ and proportions are dimensionless). (*) Values fitted considering two levels of the SARS-CoV-2 virulence.

| Symbol | Meaning | Value |
| --- | --- | --- |
| $\mu$ | Natural mortality rate | $1/(78.4 \times 365)$ |
| $\phi$ | Birth rate | $1/(78.4 \times 365)$ |
| $\varphi$ | Aging rate | $6.7 \times 10^{-6}$ |
| $\sigma_y (\sigma_o)$ | Incubation rate | $1/5.8 (1/5.8)$ |
| $\gamma_y (\gamma_o)$ | Recovery rate of asymptomatic individuals | $1/12 (1/14)$ |
| $\gamma_{1y} (\gamma_{1o})$ | Infection rate of pre-symptomatic individuals | $1/4 (1/4)$ |
| $\gamma_{2y} (\gamma_{2o})$ | Recovery rate of severe CoViD-19 individuals | $1/12 (1/21)$ |
| $\gamma_{3y} (\gamma_{3o})$ | Infection rate of mild CoViD-19 individuals | $1/13 (1/16)$ |
| $\tau^{is}$ | Time of the introduction of isolation | March 24, 2020 |
| $z_y (z_o)$ | Proportion of transmission by mild CoViD-19 individuals | $0.5 (0.2)$ |
| $\chi_y (\chi_o)$ | Proportion of remaining as asymptomatic individuals | $0.98 (0.95)$ |
| $l_y (l_o)$ | Proportion of asymptomatic individuals | * |
| $k_y (k_o)$ | Proportion of mild (non-hospitalized) CoViD-19 | * |
| $\varepsilon$ | Protection factor | * |
| $u_y (u_o)$ | Proportion in isolation | * |
| $\alpha_y (\alpha_o)$ | Additional mortality rate | * |
| $\beta_{1y} (\beta_{1o})$ | Transmission rate due to asymptomatic individuals | * |
| $\beta_{2y} (\beta_{2o})$ | Transmission rate due to pre-symptomatic individuals | * |
| $\beta_{3y} (\beta_{3o})$ | Transmission rate due to mild CoViD-19 individuals | * |

To estimate the transmission rates *β*_1*j*_, *β*_2*j*_, and *β*_3*j*_, *j* = *y, o*, the proportion in isolation *u*, and the protection factor *ε*, we use the accumulated number of severe CoViD-19 cases Ω given by

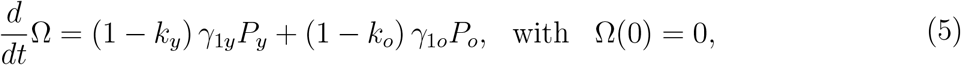

which are the exit from class *P*, and entering into class *D*.

To estimate the additional mortality rates *α*_*j*_, *j* = *y, o*, we use the number of deaths due to severe CoViD-19 cases given by Π = Π_*y*_ + Π_*o*_, where

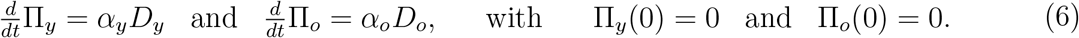

In the estimation of the additional mortality rates, we must bear in mind that the time at which new cases and deaths were registered does not have direct correspondence; instead, they are delayed by Δ days, that is, Π (*t* + Δ) = *αD*(*t*). Among exits from compartment *D* (*γ*_2*j*_, *µ*, and *α*_*j*_), we are counting only the deaths caused by severe CoViD-19.

In Appendix B, the steady-state of the system of equations in terms of fractions corresponding to equations (1), (2) and (3) was analyzed to obtain the basic reproduction number *R*_0_, which is given by equation (B.10) in Appendix B. The fractions written as 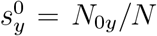 and 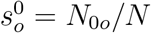 result in

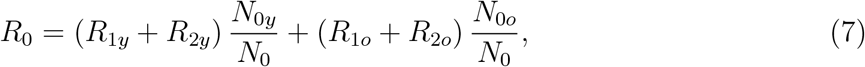

where *N*_0*y*_ and *N*_0*o*_ are the initial numbers of young and elder subpopulations with *N*_0_ = *N*_0*y*_ + *N*_0*o*_, and

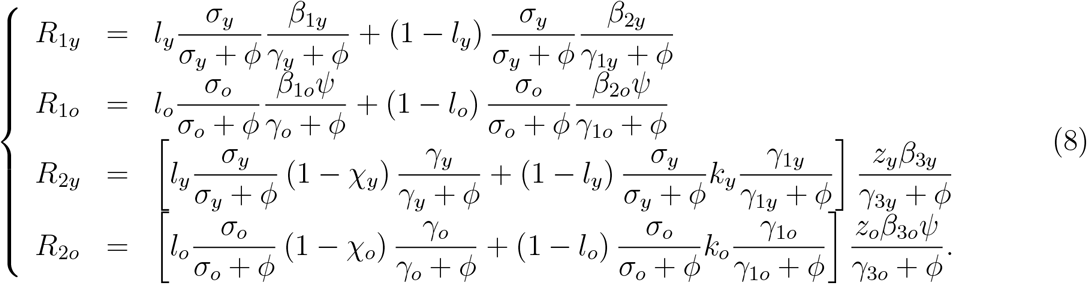

Letting *z*_*y*_ = *z*_*o*_ = 0 (*R*_2*y*_ = *R*_2*o*_ = 0), we retrieve the basic reproduction number obtained in [8].

We choose two key parameters to evaluate the trade-off between transmissibility and virulence.

1. **Transmissibility** – The force of infection *λ* given by equation (A.1) in Appendix A is the per-capita incidence rate, and *λS*_*y*_ (*λS*_*o*_) is the total number of the new cases in the young (elder) subpopulation per unit of time. Notwithstanding, the intensity of infection is proportional to the amount of virus released by infectious individuals and the capacity to infect susceptible individuals during contacts between them. The transmission rates carry on this fact besides other characteristics not considered here (social network, movement, demography, genetic, nutritional and health conditions, etc.). For this reason, we assume that the basic reproduction number *R*_0_ calculated by the estimated transmission rates measures the transmissibility of SARS-CoV-2.
2. **Virulence** – Enhancing the virus’s capacity to infect target cells, an increased number of cells are infected. Consequently, the possibility of manifesting mild or severe disease increases, increasing the symptomatic CoViD-19 cases. For this reason, we translate the virulence with the ratio between asymptomatic and symptomatic individuals.

Notice that another effect of higher virulence is the increased amount of virus released by infectious individuals in the environment, which must increase the transmission. This fact is currently accepted for CoViD-19 epidemic [6]. We consider two broadly separated values for the proportion of asymptomatic individuals and estimate the transmission rates to calculate the basic reproduction number *R*_0_.

## 3 Results

At the beginning of the epidemic, we have two data sets: Severe CoViD-19 cases (those in hospitals were tested and confirmed) and deaths. We study the transmissibility and virulence of SARS-CoV-2 considering the data collection from São Paulo State, Brazil [9]. Due to the lack of mass testing (PCR and serology) when SARS-CoV-2 transmission begins, the available severe CoViD-19 curve *D* is considered the epidemic curve.

To evaluate the trade-off between transmissibility and virulence, we assumed that the basic reproduction number *R*_0_ measures the transmissibility, and the proportions of asymptomatic individuals *l*_*y*_ and *l*_*o*_ assess the virulence. Letting *l*_*y*_ = *l*_*o*_, we consider two values: *l*_1_ = 0.8 (lower virulence, with the ratio between asymptomatic and symptomatic individuals being 4 : 1) and *l*_2_ = 0.2 (higher virulence, with the ratio 1 : 4).

To estimate the transmission rates, the proportion in isolation, and the protective factor (*x* stands for one of these parameters), we calculate

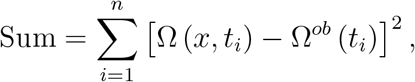

where Ω (*x, t*_*i*_) is the accumulated severe CoViD-19 cases calculated from equation (5) and Ω^*ob*^ (*t*_*i*_) is the accumulated severe CoViD-19 registered cases at day *t*_*i*_. We search for the value of *x* minimizing the Sum. To evaluate Ω, the numerical solutions of equations (1), (2) and (3) are obtained using the initial conditions given by equation (A.4) in Appendix A. To estimate the additional mortality rates, we substitute Ω (*x, t*_*i*_) by Π (*α, t*_*i*_) given by equation (6) and Ω^*ob*^ (*t*_*i*_) by Π^*ob*^ (*t*_*i*_).

The observed data from February 26 to November 31 [9] is partitioned into two sets – The first set is used to estimate the model parameters (input set), and the estimated model is then confronted with the second set to assess its prediction ability (test set) [10]. Hence, we estimate the model parameters using severe CoViD-19 (Ω^*ob*^) and deaths (Π^*ob*^) data from February 26 to May 13, 2020 (input data set). Then, we use the estimated parameters to predict the epidemic under interventions (quarantine and relaxation) from May 14 to November 31 (test data set) and compare the outcomes with the observed data.

We estimate the model parameters using the data from February 26 to May 13 following the estimation procedure presented in [7]. For higher virulence (*l*_2_ = 0.2), we obtain (see Figure 1 below):

**Figure 1:**
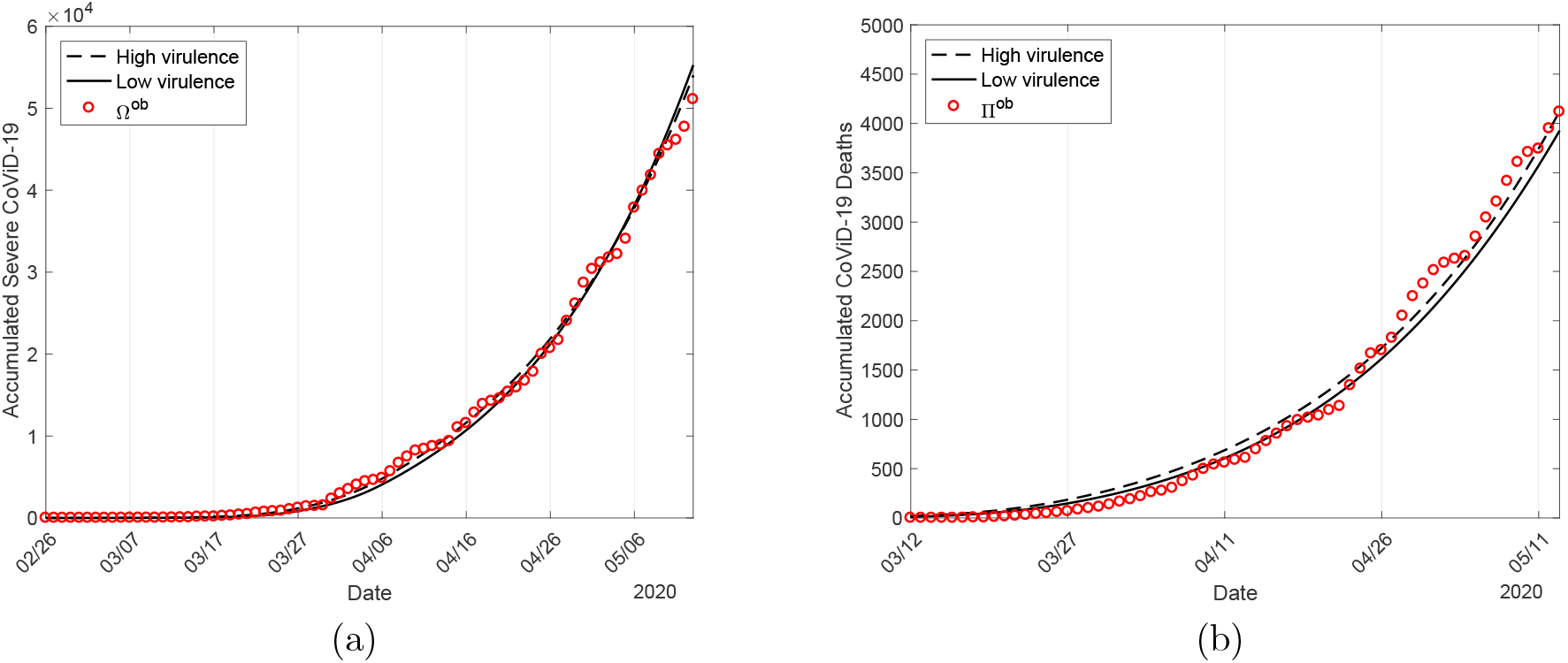
The estimated accumulated curve Ω and the observed data Ω^*ob*^ (a), and the accumulated deaths Π and the observed Π^*ob*^ (b) for the higher (continuous line) and lower (dashed line) virulence. Estimations were done considering data from São Paulo State.

1. **Transmission rates** (from February 26 to April 3) – Estimated values are *β*_*y*_ = 0.69 and *β*_*o*_ = 0.79 (both in *days*^−1^), resulting in the basic reproduction number *R*_0_ = 7.02. When the CoViD-19 epidemic was initiated, we did not have any control mechanisms, which is characterized as the natural epidemic. The effect of isolation on March 24 appears around 9 days later.
2. **Proportion in partial quarantine** (from March 24 to April 13) – Estimated value is *k* = 0.48. The effect of adopting protective measures on April 4 appears around 9 days later.
3. **Protective measures** (from April 4 to May 13) – Estimated value is *ε* = 0.53.
4. **Additional mortality rates** (from March 16 to May 28) – Estimated values are *α*_*y*_ = 0.0018 and *α*_*o*_ = 0.0071 (both in *days*^−1^) fixing Δ = 15 *days*. The first death due to CoViD-19 was on March 16, and we considered 15 more observed data.

The estimated parameters are summarized in Table 3. For lower virulence (*l*_2_ = 0.8), we transport the estimated parameters from [7]. The period from February 26 to May 13 characterizes the beginning of the epidemic under partial quarantine (isolation) and protective measures adopted by individuals. Based on the two virulence levels’ estimated parameters considering the input data set, we study the epidemic’s prediction ability during the following test data set.

**Table 3:** The estimated parameters (*j* = *y, o*) considering two SARS-CoV-2 virulence levels (rates in *days*^−1^ and proportions are dimensionless).

| Parameter | Low ($l_1 = 0.8$) | High ($l_2 = 0.2$) |
| --- | --- | --- |
| $k_y (k_o)$ | 0.92 (0.75) | 0.94 (0.78) |
| $\varepsilon$ | 0.5 | 0.53 |
| $u_y (u_o)$ | 0.53 (0.53) | 0.48 (0.48) |
| $\alpha_y (\alpha_o)$ | 0.00185 (0.0071) | 0.00185 (0.0071) |
| $\beta_{1y} (\beta_{1o})$ | 0.78 (0.90) | 0.69 (0.79) |
| $\beta_{2y} (\beta_{2o})$ | 0.78 (0.90) | 0.69 (0.79) |
| $\beta_{3y} (\beta_{3o})$ | 0.78 (0.90) | 0.69 (0.79) |
| $R_0$ | 9.24 | 7.02 |

Figure 1 shows the estimated curve of accumulated CoViD-19 cases Ω and the observed data Ω^*ob*^ (a), and the accumulated deaths Π and the observed Π^*ob*^ (b) for the higher (continuous line) and lower (dashed line) virulence. Estimations were done considering São Paulo State’s data from February 26 to May 13, 2020, for Ω, and from March 16 to May 28 for Π.

The natural epidemic is characterized by the estimated transmission rates from February 26 to April 3. Since then, the observed data correspond to the epidemic with interventions (quarantine and relaxation). Figure 2 shows the extended accumulated Covid-19 cases Ω (a) and the corresponding natural epidemic curves *D* (b) for the higher (continuous line) and lower (dashed line) virulence.

**Figure 2:**
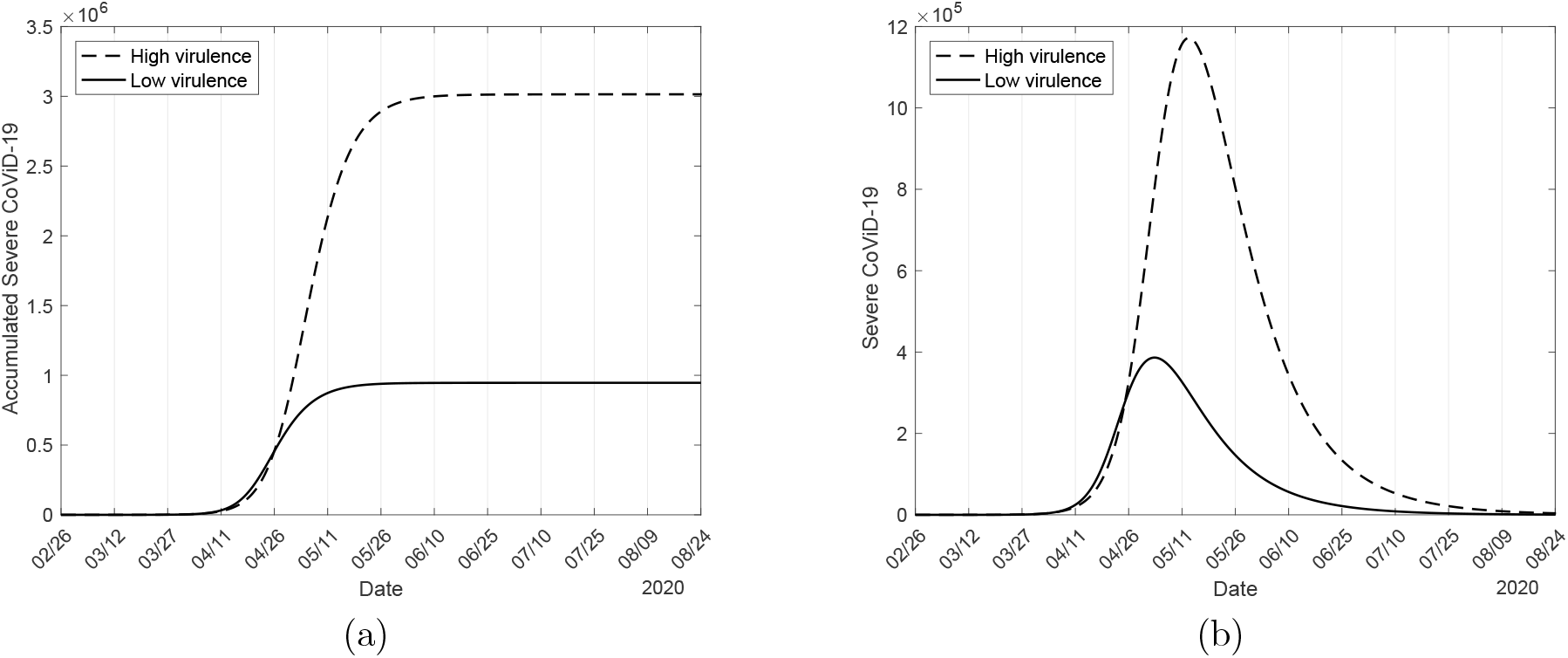
The extended accumulated Covid-19 cases Ω (a) and the corresponding natural epidemic curves *D* (b) for the higher (continuous line) and lower (dashed line) virulence.

Figure 2(a) shows that at the end of the epidemic’s first wave, Ω approaches the asymptote (plateaux) 9.46 × 10^5^ for lower virulence and 3.01 × 10^6^ for higher virulence. The curve of deaths Π (not shown) has the sigmoid shape as Ω and approaches the asymptote 5.73 × 10^4^ for lower virulence, and 1.95 × 10^5^ for higher virulence. Figure 2(b) shows that the natural epidemic’s peak is 3.86 × 10^5^, occurring on May 3, 2020, for lower virulence, and 1.17 × 10^6^ occurring on May 12 for higher virulence. In the natural epidemic (Figures 1 until April 3, and 2), the estimated curves of Ω and Π are quite the same in the initial phase for lower and higher virulence strains; however, the latter approaches to asymptotic values increased by around 330%. The peak of the higher virulent SARS-CoV-2 epidemic curve is increased by around 300% from the lower one; hence we take the peaks of curves and not the asymptotic values to compare epidemic outcomes.

Let us evaluate the estimated parameters’ prediction ability considering two virulence levels on the epidemic under interventions in São Paulo State. Besides the input data set, we include the test data set and show the observed data Ω^*ob*^ and Π^*ob*^ from February 26 to November 31. The model is solved numerically during the input and test data sets using the estimated parameters given in Table 3. The resulting epidemiological scenarios correspond to the quarantine epidemic and are shown in Figures 3 to 5.

**Figure 3:**
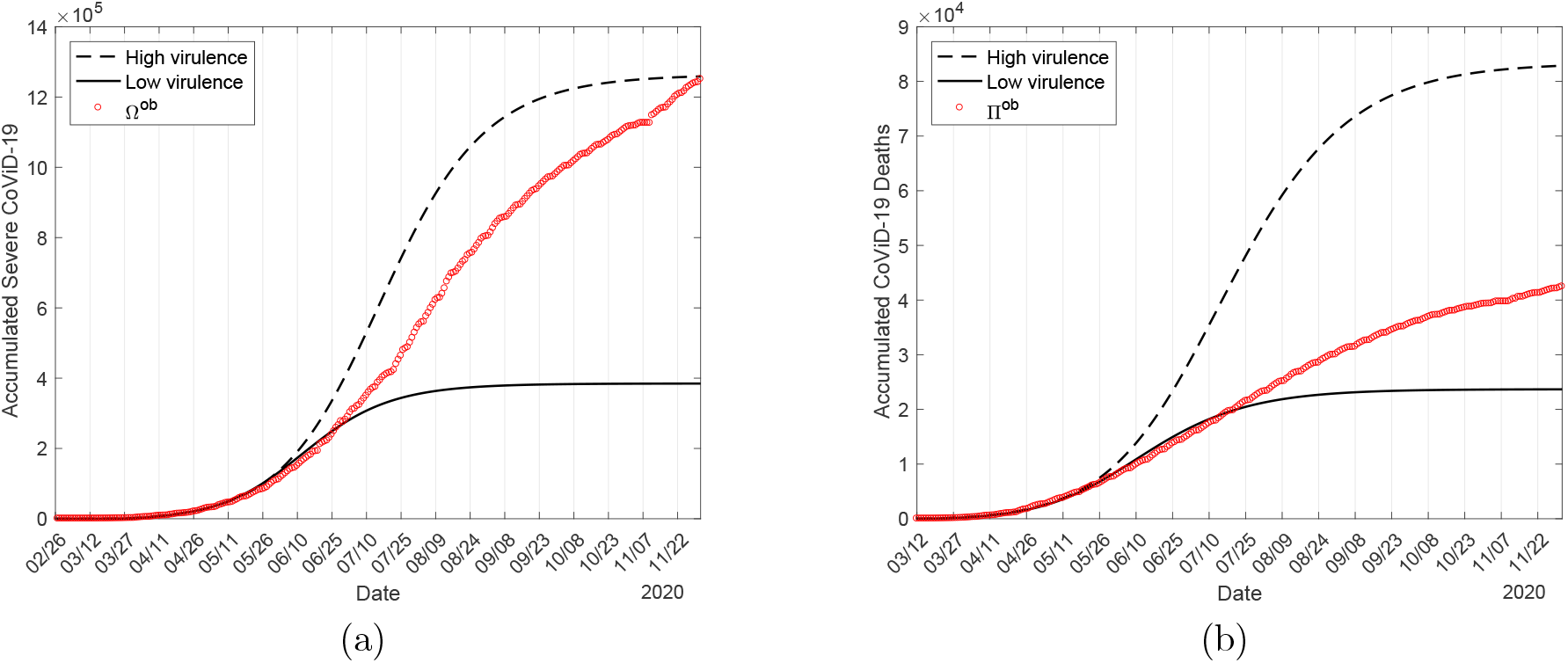
The extended curves of the accumulated CoViD-19 cases Ω (a) and deaths Ω (b) for higher (continuous line) and lower (dashed line) virulence. The curves correspond to the epidemic under isolation.

Figure 3 shows the curves of the accumulated CoViD-19 cases Ω (a) and deaths Π (b) for higher (continuous line) and lower (dashed line) virulence. Notice that Figure 3 is the extension of Figure 1. The lower and higher virulence estimations’ prediction ability is analyzed below by comparing the epidemic outcomes with the observed data Ω^*ob*^ and Π^*ob*^.

From Figure 3, for lower virulence, the accumulated CoViD-19 cases at the end of the epidemic under isolation’s first wave Ω and deaths Π are 3.85 × 10^5^ (41%) and 2.37 × 10^4^ (41.3%), and for higher virulence, they are 1.26 × 10^6^ (41.9%) and 8.29 × 10^4^ (42.5%). The percentage between parentheses is the ratio to the natural epidemic. The partial quarantine decreased by around 40% of the severe CoViD-19 cases and deaths from the natural epidemic in lower and higher virulence. For the higher virulence, the asymptotic values of Ω and Π increased by around 340% from the lower virulence, close to the natural epidemic’s increase.

Figure 4 shows the curves of asymptomatic (*A*) and pre-symptomatic (*P*) individuals (a), and the estimated curves of mild (*M*) and severe (*D*) CoViD-19 cases (b) for higher (continuous line) and lower (dashed line) virulence. Figure 4 also shows the curve of carriers capable of transmitting the virus (*A* + *P*) and the curve of apparent CoViD-19 (*M* + *D*).

**Figure 4:**
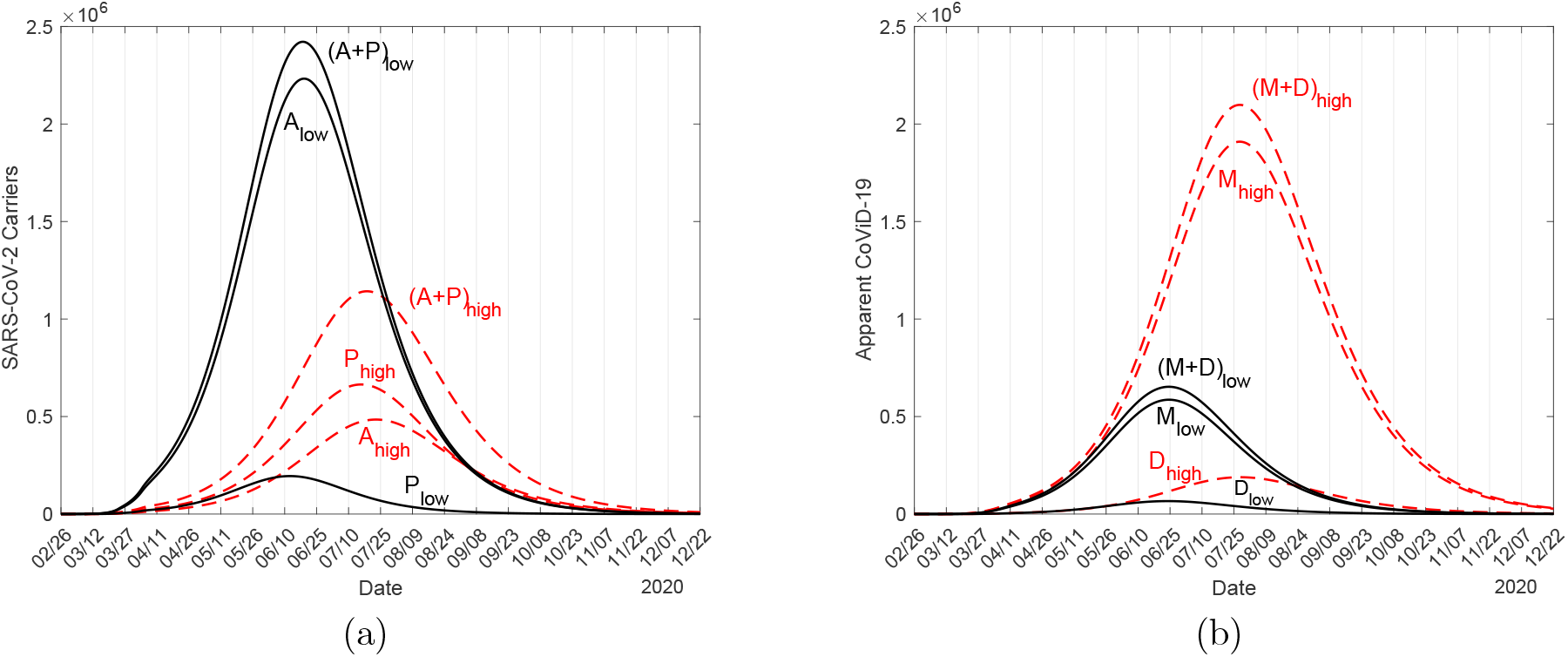
The estimated curves of asymptomatic (*A*) and pre-symptomatic (*P*) individuals (a), and the estimated curves of mild (*M*) and severe (*D*) CoViD-19 cases (b) for higher (continuous line) and lower (dashed line) virulence.

Figure 4 illustrates the impact of control mechanisms on flattening the natural epidemic curve. For lower virulence, the peaks of asymptomatic (*A*), pre-symptomatic (*P*), mild (*M*), and severe (*D*) CoViD-19 are 2.23 × 10^6^, 1.95 × 10^5^, 5.86 × 10^5^, and 0.66 × 10^5^, and for higher virulence, the peaks are 4.85 × 10^5^, 6.64 × 10^5^, 1.91 × 10^6^, and 1.89 × 10^5^. (For the lower virulence, the peak of *A* is increased by 460%, and the peaks of *P, M*, and *D* are reduced by 340%, 326%, and 286% from the higher virulence.) The asymptomatic and pre-symptomatic individuals are carriers, which difficult the control of infection. Comparing the lower and higher virulence, the ratio between the peaks of carriers (*A* + *P*) is 212%, and the ratio between peaks of apparent CoViD-19 (*M* + *D*) is 31%.

Figure 5 shows the curves of transmitting SARS-CoV-2 individuals (*A* + *P* + *z*_*y*_*M*_*y*_ + *z*_*o*_*M*_*o*_) (a), and the curves of harboring SARS-CoV-2 individuals (*E, A, P, M, D*) (b) for higher (continuous line) and lower (dashed line) virulence.

**Figure 5:**
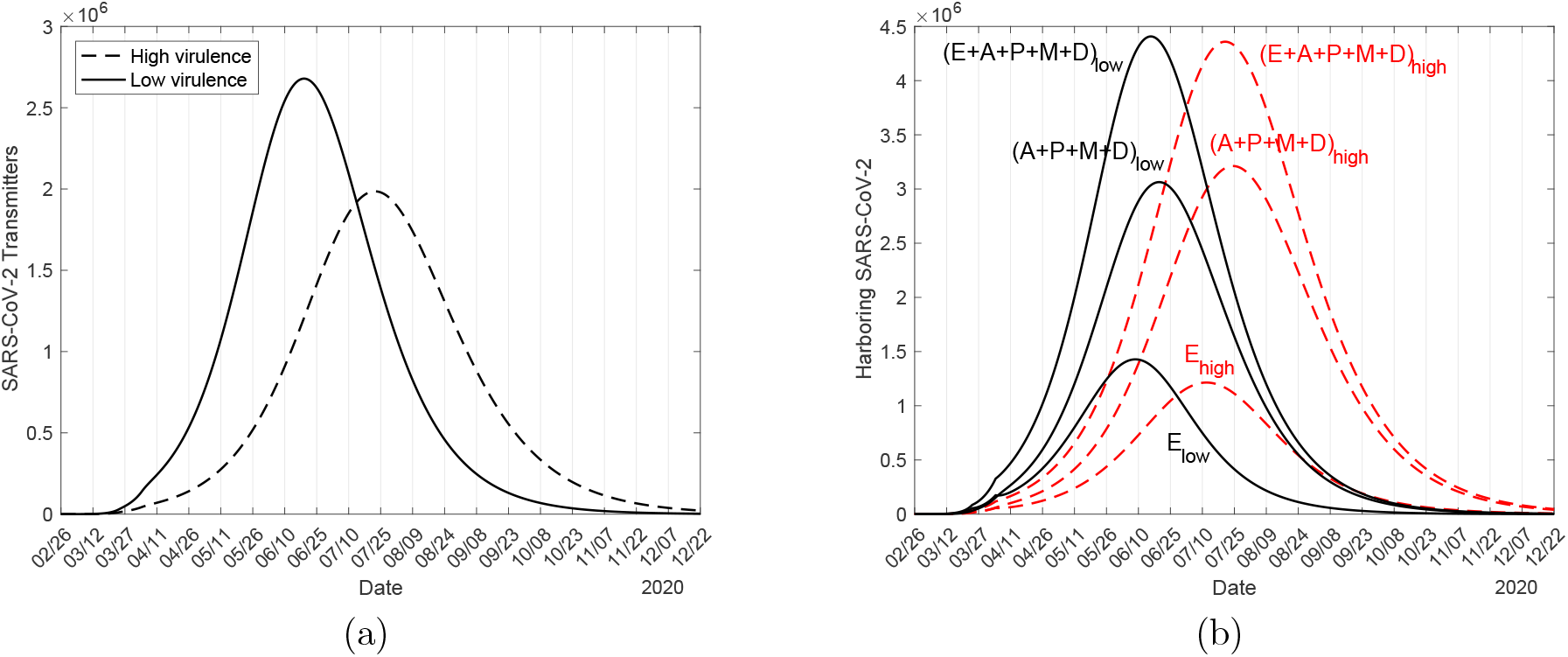
The curves of transmitting SARS-CoV-2 individuals (a), and the curves of harboring SARS-CoV-2 individuals (b) for higher (continuous line) and lower (dashed line) virulence.

From Figure 5(a), for lower virulence, the peaks of transmitting SARS-CoV-2 individuals (*A* +*P* +*z*_*y*_*M*_*y*_ +*z*_*o*_*M*_*o*_) for lower and higher virulence are 2.68 × 10^6^ and 1.99 × 10^6^, occurring on June 19 and July 21. The ratio between peaks of all infectious individuals with lower and higher virulence is 135%. From Figure 5(b), for lower virulence, the peaks of exposed (*E*), releasing virus (*A* + *P* + *M* + *D*), and harboring virus (*E* + *A* + *P* + *M* + *D*) individuals are 1.43 × 10^6^, 3.06 × 10^6^, and 4.41 × 10^6^, and for higher virulence, the peaks are 1.21 × 10^6^ (85%), 3.21 × 10^6^ (105%), and 4.36 *×* 10^6^ (99%). The percentage between parentheses is the ratio to the lower virulence. The individuals releasing the virus are slightly higher in a higher virulence, but all individuals harboring the virus are practically equal in lower and higher virulence transmissions. Notice that exposed individuals (the new infection cases) are higher in lower virulence due to a higher *R*_0_. The peaks occur close to those observed in Figure 5(a).

Figures 3-5 showed that the enhanced virulence increased the deaths due to CoViD-19 by around 300%; however, the transmissibility measured by *R*_0_ was reduced by around 76%. The higher virulence increased by around 300% the number of mild and severe CoViD-19 cases, potentially more transmissible but in isolation; however, the number of asymptomatic, presymptomatic, and a fraction of mild CoViD-19 individuals transmitting the virus is reduced by 74%. The SARS-CoV-2 transmission decreases in the populational point of view, and the number of deaths rises as the virulence increases. Initially, the model variables (compartments) have quite the same values, but the peaks of the more virulent SARS-CoV-2 occur around one month later. In particular, the number of deaths in the more virulent epidemic is three times higher than the less one, although both strains presented the same fatalities during the first three months.

Let us assess the prediction ability by comparing the observed data with the lower and higher virulence epidemiological scenarios. Remember that the lower and higher virulent SARS-CoV-2, considering the input data set comprising the quarantine period, provided quite similar fitted Ω and Π curves (see Figure 1). However, from Figure 3, the lower virulent epidemic fits very well the subset of test data from May 13 to June 30 (Ω) or July 20 (Π), showing that quarantine lasted until that date. (The first observed point detaching from the lower virulence curve Π occurs on July 21, while the severe CoViD-19 curve Ω separates on July 1, 20 days earlier. In the estimation of mortality rates *α*_*y*_ and *α*_*o*_, we used Δ = 15 days in Π (*t* + Δ) = *αD*(*t*), not Ω.) São Paulo State initiated the relaxation by mid-June 2020, with the effect on the epidemic curve *D* appearing 9 days later. Hence, the subset of test data since July 1 must be described by a model incorporating intermittent pulses of release [8], which is not dealt with here. Therefore, the less virulent SARS-CoV-2 transmission explains better the CoViD-19 epidemic until the beginning of the relaxation.

The relation Π (*t* + Δ) = *αD*(*t*) does not sustain due to the mass test including mild and eventually asymptomatic and pre-symptomatic individuals in the observed data. This fact is apparent in Figure 3, where the registered SARS-CoV-2 infections increase more quickly than the observed deaths. The supplementary deaths since July 21 can be explained by the beginning of the relaxation 20 days earlier. Moreover, the registered number of deaths in São Paulo State is 58873 on February 25, 2021, which is higher than 57300 predicted by the low virulent natural epidemic (see underneath Figure 2). However, São Paulo State is still in partial quarantine. Hence, based on the model, the supplementary deaths can not be explained only by the relaxation but by the circulation of more virulent variant(s) of the original SARS-CoV-2.

## 4 Discussion and conclusion

The persistent transmission of SARS-CoV-2 should result in a high number of mutations and variants. In this pool of variants, some acquired enhanced ability to infect cells, increasing the risk of death. Moreover, more viruses will be released in the environment when more cells are infected, increasing the transmission. However, the severe symptomatic individuals are isolated in hospitals, and mild cases are detected by test and advised to isolate. Contrarily, the asymptomatic and pre-symptomatic individuals (carriers) are not seen and circulate freely. Additionally, the carriers are in close contact with other susceptible individuals, contrarily to individuals manifesting any suspicious symptoms (not necessarily CoViD-19). This fact was explained by the model, showing that higher virulence causes increased deaths but is less transmissible – By the fact that *R*_0_ is lower, the epidemic curve increases more slowly, but the curve of deaths increases much more quickly than the less virulent one.

In the first months of the CoViD-19 outbreak, the absence of a mass test against SARS-CoV-2 infection and clinical follow-up of infected individuals did not permit the discrimination between asymptomatic and pre-symptomatic individuals. Considering this initial period, we explained the CoViD-19 data from São Paulo State using two broadly separated values for the ratio between asymptomatic and symptomatic individuals (4 : 1 and 1 : 4). For each proportion *l*, we estimated the transmission rates and calculated the basic reproduction number *R*_0_.

Figures 3-5 showed two broadly separated virulence to explain CoViD-19 data from São Paulo State. The partial quarantine and protective measures flattened the epidemic curve in São Paulo State, but the transmission was maintained at a relatively higher level (the effective reproduction number reached one around 80 days later [7]). This epidemiological scenario can be explained by spreading a less virulent original SARS-CoV-2 or a pool of less virulent variants dominating during the quarantine period. However, since July 1, the observed number of deaths was systematically above the estimated curve of Π considering quarantine only. The effects of relaxation initiated in São Paulo State appeared on July 1, but it was not fully implemented. We do not have a realistic assessment of the relaxation neither the transmission of SARS-CoV-2 among the isolated individuals (in [7], a decreased transmission in the isolated population is allowed). At the end of February 2021, São Paulo State still maintained part of the population in isolation, but 58873 deaths on February 25, 2021, surpassed 57300 predicted by the natural epidemic under quarantine. This higher number of deaths under isolation may be a consequence of the appearance of more virulent variants (but not so high as *l* = 0.2 shown in Figures 1-5) during the one-year SARS-CoV-2 transmission. Hence, based on the model’s analysis, we can assume that virulent(s) SARS-CoV-2 can be found in the current pool of mutants, resulting in a continuously increasing number of severe CoViD-2 and deaths. However, more detection of higher virulence severe cases and deaths is synonymous with more transmissibility is a wrong statement according to the model.

The epidemiological scenario in Amazonas State (Brazil) can shed more light on the virulent variants’ role in association with relaxation. In Amazonas State, the occurrence of deaths in January-February, 2021, is higher than that occurred during all 2020 [11]. This increased number can not be explained only by the partial relaxation once a significant number of deaths caused by the P.1 variant was found. Hence, Amazonas State is another example of a pool containing high virulent SARS-CoV-2 transmission.

Finally, during the epidemic, the fast mutations in RNA-virus can result in a pool of variants. This pool can behave on average with lower or higher virulence than the original SARS-CoV-2. In the short-run epidemic, if a pool of variants acts on average as a high (low) virulent strain, the number of severe cases and deaths increases (decreases), but the transmission is decreased (increased). However, this trade-off between transmissibility and virulence shows that, in the long-run epidemic, lower virulence with higher transmissible variant(s) will prevail. Therefore, in the short-run epidemic control, strict isolation of symptomatic and perhaps suspicious individuals must be adopted to avoid high virulent SARS-CoV-2 transmission causing more death risk.

## Data Availability

The data that support the findings of this study are openly available in "SP contra o novo coronavirus (Boletim completo)" at https://www.seade.gov.br/coronavirus/.

## Availability of data and material

The data that support the findings of this study are openly available in SP contra o novo coronavírus (Boletim completo) at https://www.seade.gov.br/coronavirus/.

## Author contributions

Hyun Mo Yang: Conceptualization, Methodology, Formal analysis, Writing - Original draft preparation, Validation, Supervision. Luis Pedro Lombardi Junior: Software, Data Curation, Visualization, Validation. Ariana Campos Yang: Conceptualization, Validation, Investigation.

## A The SQEAPMDR model

One of the main aspects of CoViD-19 is increased fatality in the elder subpopulation. For this reason, a population is divided into two groups, composed of young (60 years old or less, denoted by subscript *y*) and elder (60 years old or more, denoted by subscript *o*) subpopulations. This community’s vital dynamic is described by the per-capita rates of birth (*ϕ*) and mortality (*µ*), and *φ* is the aging rate, that is, the flow from young subpopulation *y* to elder subpopulation *o*. Another aspect is the presence of the pre-symptomatic individuals, that is, individuals without symptoms transmitting SARS-CoV-2 before the onset of the disease [12].

Hence, for each subpopulation *j* (*j* = *y, o*), individuals are divided into seven compartments: susceptible *S*_*j*_, isolated (quarantine) *Q*_*j*_, exposed and incubating *E*_*j*_, asymptomatic *A*_*j*_, pre-symptomatic (or pre-diseased) individuals *P*_*j*_, symptomatic individuals with mild CoViD-19 *M*_*j*_, and severe CoViD-19 *D*_*j*_. However, all young and elder individuals in classes *A*_*j*_, *M*_*j*_, and *D*_*j*_ enter into the same recovered class *R* (this is the 8^*th*^ class, but common to both subpopulations). Hence, the SQEAPMDR model has 15 compartments.

The natural history of CoViD-19 is the same for young (*j* = *y*) and elder (*j* = *o*) subpopulations. We assume that individuals in the asymptomatic (*A*_*j*_), pre-diseased (*P*_*j*_), and a fraction *z*_*j*_ of mild CoViD-19 (*M*_*j*_) classes are transmitting the virus. Other infected classes ((1 − *z*_*j*_) *M*_*j*_ and *D*_*j*_) are under voluntary or forced isolation. Susceptible individuals are infected at a rate *λ*_*j*_*S*_*j*_ (known as the mass action law [13]), where *λ*_*j*_ is the per-capita incidence rate (or force of infection) defined by *λ*_*j*_ = *λ* (*δ*_*jy*_ + *ψδ*_*jo*_), with *λ* being

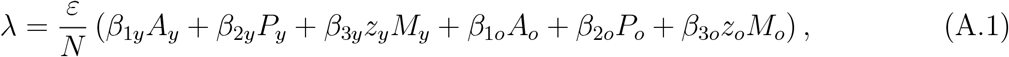

where *δ*_*ij*_ is the Kronecker delta, with *δ*_*ij*_ = 1 if *i* = *j*, and 0, if *i* ≠ *j*; and *β*_1*j*_, *β*_2*j*_ and *β*_3*j*_ are the transmission rates, that is, the rates at which a virus encounters a susceptible people and infects him/her. In [8], a particular model was analyzed letting *z*_*y*_ = *z*_*o*_ = 0 and *χ*_*y*_ = *χ*_*o*_ = 1.

Susceptible individuals are infected at a rate *λ*_*j*_ and enter into class *E*_*j*_. After an average period 1*/σ*_*j*_ in class *E*_*j*_, where *σ*_*j*_ is the incubation rate, exposed individuals enter into the asymptomatic class *A*_*j*_ (with probability *l*_*j*_) or pre-diseased class *P*_*j*_ (with probability 1 − *l*_*j*_). After an average period 1*/γ*_*j*_ in class *A*_*j*_, where *γ*_*j*_ is the recovery rate of asymptomatic individuals, asymptomatic individuals acquire immunity (recovered) and enter into recovered class *R*. Possibly asymptomatic individuals can manifest symptoms at the end of this period, and a fraction 1 − *χ*_*j*_ enters into mild CoViD-19 class *M*_*j*_. For symptomatic individuals, after an average period 1*/γ*_1*j*_ in class *P*_*j*_, where *γ*_1*j*_ is the infection rate of pre-diseased individuals, pre-diseased individuals enter into severe CoViD-19 class *D*_*j*_ (with probability 1 − *k*_*j*_) or mild CoViD-19 class *M*_*j*_ (with probability *k*_*j*_). Individuals in class *D*_*j*_ acquire immunity after a period 1*/γ*_2*j*_, where *γ*_2*j*_ is the recovery rate of severe CoViD-19, and enter into recovered class *R* or die under the disease-induced (additional) mortality rate *α*_*j*_. Individuals in mild CoViD-19 class *M*_*j*_ acquire immunity after a period 1*/γ*_3*j*_, where *γ*_3*j*_ is the recovery rate of mild CoViD-19, and enter into recovered class *R*.

The dynamic equations for *E, A, P, M, D*, and *R* are obtained through the balance between inflow and outflow in each compartment (see equations (2) and (3) in the main text). For the equations corresponding to *S* and *Q*, we consider a unique pulse in isolation at time *t* = *τ*, described by *u*_*j*_*S*_*j*_*δ* (*t* − *τ*), with *j* = *y, o*. The fraction of persons in isolation is *u*_*j*_ and *δ* (*x*) is the Dirac delta function, that is, *δ* (*x*) =∞, if *x* = 0, otherwise, *δ* (*x*) = 0, with 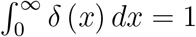. The isolation is translated as

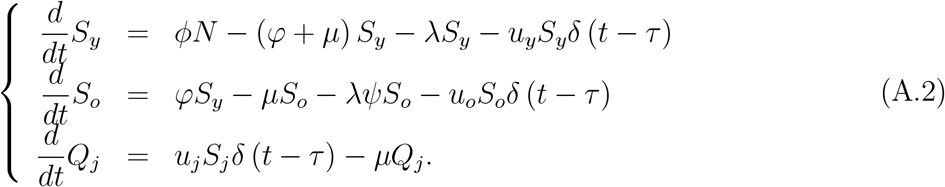

These equations are simulated permitting intermittent interventions to the boundary conditions.

The initial conditions (simulation time *t* = 0) supplied to the system of equations (1), (2), and (3) are, for *j* = *y, o*,

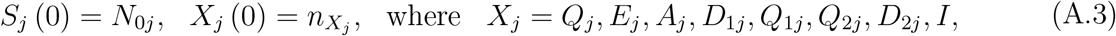

where 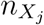 is a non-negative number. For instance, 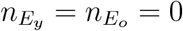 means that there is not any exposed person (young and elder) at the beginning of the epidemic. In this paper, we use

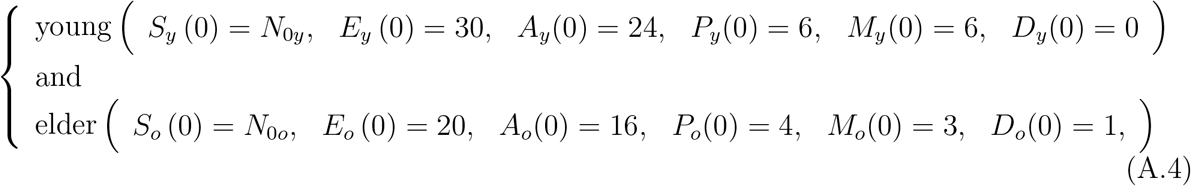

plus *Q*_*y*_ (0) = *Q*_*o*_ (0) = *R*(0) = 0, where *t* = 0 corresponds to the calendar time when the first case was confirmed (February 26 for São Paulo State). For São Paulo State, *N*_0*y*_ = 37.8 million and *N*_0*o*_ = 6.8 million. (See [7] for details in the initial conditions’ setup.)

The isolation implemented at *τ* = 27 (corresponding to calendar time March 24, 2020) is described by the boundary conditions

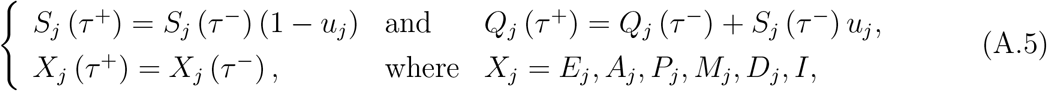

with *τ*^−^ = lim_*t*→*τ*_ *t* (for *t* < *τ*), and *τ* ^+^ = lim_*τ*←*t*_ *t* (for *t* > *τ*).

## B The steady-state analysis of the SQEAPMDR model

The system of equations (1), (2) and (3) does not reach steady-state (time varying population). However, this system in term of fractions attains steady-state. Defining the fraction *x*_*j*_ = *X*_*j*_*/N*, for *j* = *y, o*, with *X*_*j*_ = {*S*_*j*_, *Q*_*j*_, *E*_*j*_, *A*_*j*_, *P*_*j*_, *M*_*j*_, *D*_*j*_, *R*}, we have

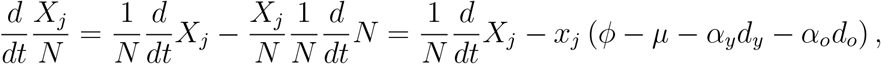

using equation (4), and the system of equations (1), (2) and (3) in terms of fractions become, for susceptible individuals,

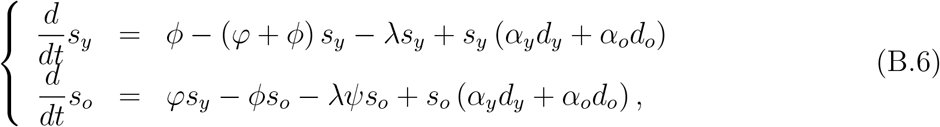

for isolated and infected individuals,

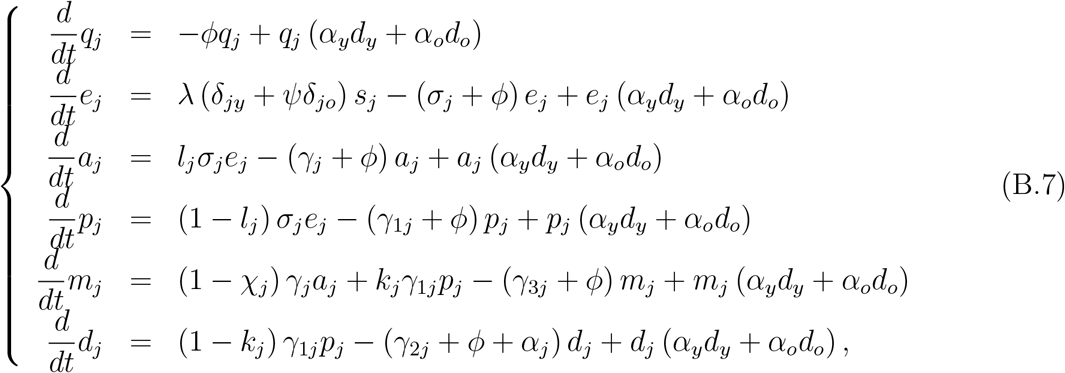

and for recovered individuals,

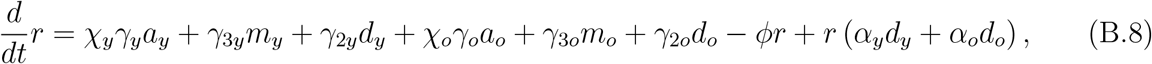

where *λ* is the force of infection given by equation (A.1) re-written as

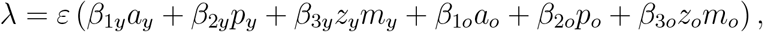

and

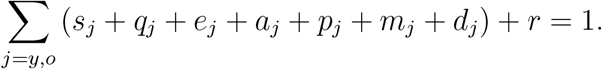

This new system of equations has a steady-state, that is, the number of individuals in all classes varies with time. However, their fractions attain a steady-state (the sum of derivatives of all classes is zero).

The trivial (disease-free) equilibrium point *P* ^0^ of the new system of equations (B.6), (B.7) and (B.8) is given by

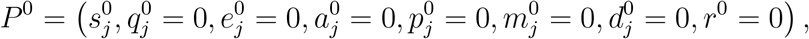

for *j* = *y* and *o*, where

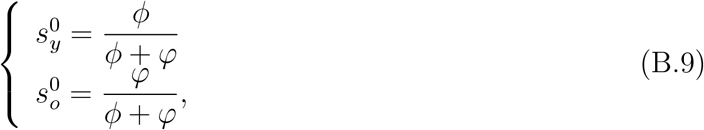

with 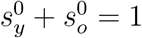.

Let us assess the stability of *P* ^0^ by applying the next generation matrix theory considering the vector of variables *x* = (*e*_*y*_, *a*_*y*_, *p*_*y*_, *m*_*y*_, *e*_*o*_, *a*_*o*_, *p*_*o*_, *m*_*o*_) [14]. We apply method proposed in [15] and proved in [16]. To obtain the basic reproduction number, diagonal matrix *V* is considered. Hence, the vectors *f* and *v* are

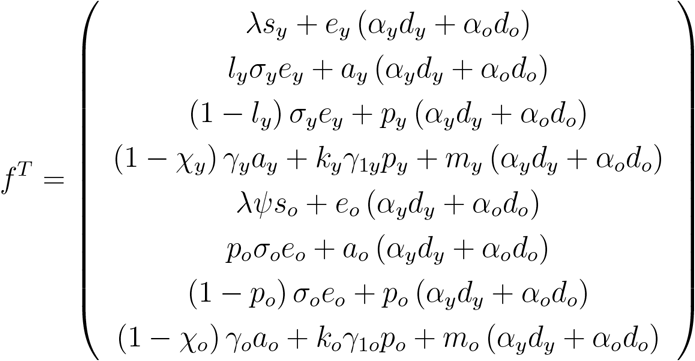

and

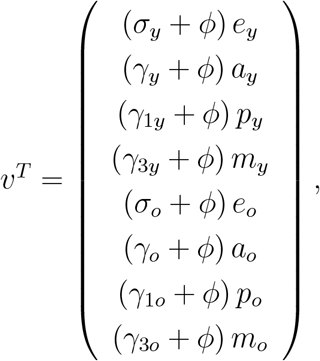

where the superscript *T* stands for the transposition of a matrix, from which we obtain the matrices *F* and *V* (see [14]) evaluated at the trivial equilibrium *P* ^0^, which were omitted. The next generation matrix *FV* ^−1^ is

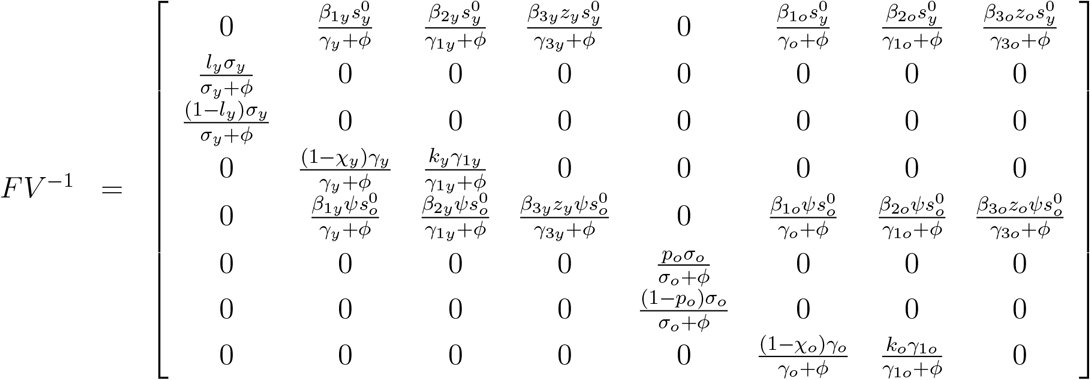

and the characteristic equation corresponding to *FV* ^−1^ is

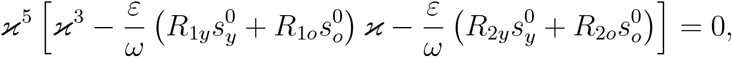

with the basic reproduction number *R*_0_ being given by

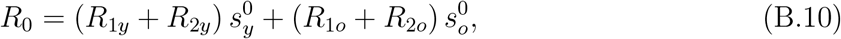

where the initial fractions 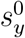 and 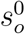 are given by equation (B.9), and the partial basic reproduction numbers *R*_1*y*_, *R*_2*y*_, *R*_1*o*_, and *R*_2*o*_ are given by equation (8) in the main text. The spectral radius *ρ* (*FV* ^−1^) is the biggest solution of a third-degree polynomial, not easy to evaluate. The procedure proposed in [15] allows us to obtain the threshold *R*_0_ as the sum of coefficients of the characteristic equation. Hence, the trivial equilibrium point *P* ^0^ is locally asymptotically stable if *R*_0_ < 1.

